# Holistic AI analysis of hybrid cardiac perfusion images for mortality prediction

**DOI:** 10.1101/2024.04.23.24305735

**Authors:** Anna M Michalowska, Wenhao Zhang, Aakash Shanbhag, Robert JH Miller, Mark Lemley, Giselle Ramirez, Mikolaj Buchwald, Aditya Killekar, Paul B Kavanagh, Attila Feher, Edward J Miller, Andrew J Einstein, Terrence D Ruddy, Joanna X Liang, Valerie Builoff, David Ouyang, Daniel S Berman, Damini Dey, Piotr J Slomka

## Abstract

**Background:** While low-dose computed tomography scans are traditionally used for attenuation correction in hybrid myocardial perfusion imaging (MPI), they also contain additional anatomic and pathologic information not utilized in clinical assessment. We seek to uncover the full potential of these scans utilizing a holistic artificial intelligence (AI)-driven image framework for image assessment.

**Methods:** Patients with SPECT/CT MPI from 4 REFINE SPECT registry sites were studied. A multi-structure model segmented 33 structures and quantified 15 radiomics features for each on CT attenuation correction (CTAC) scans. Coronary artery calcium and epicardial adipose tissue scores were obtained from separate deep-learning models. Normal standard quantitative MPI features were derived by clinical software. Extreme Gradient Boosting derived all-cause mortality risk scores from SPECT, CT, stress test, and clinical features utilizing a 10-fold cross-validation regimen to separate training from testing data. The performance of the models for the prediction of all-cause mortality was evaluated using area under the receiver-operating characteristic curves (AUCs).

**Results:** Of 10,480 patients, 5,745 (54.8%) were male, and median age was 65 (interquartile range [IQR] 57-73) years. During the median follow-up of 2.9 years (1.6-4.0), 651 (6.2%) patients died. The AUC for mortality prediction of the model (combining CTAC, MPI, and clinical data) was 0.80 (95% confidence interval [0.74-0.87]), which was higher than that of an AI CTAC model (0.78 [0.71-0.85]), and AI hybrid model (0.79 [0.72-0.86]) incorporating CTAC and MPI data (p<0.001 for all).

**Conclusion:** In patients with normal perfusion, the comprehensive model (0.76 [0.65-0.86]) had significantly better performance than the AI CTAC (0.72 [0.61-0.83]) and AI hybrid (0.73 [0.62-0.84]) models (p<0.001, for all).CTAC significantly enhances AI risk stratification with MPI SPECT/CT beyond its primary role - attenuation correction. A comprehensive multimodality approach can significantly improve mortality prediction compared to MPI information alone in patients undergoing cardiac SPECT/CT.

## 1. Introduction

Myocardial perfusion scintigraphy is widely used for the evaluation of coronary artery disease (CAD), with over 15-20 million scans performed worldwide.^1,2^ During myocardial perfusion imaging (MPI), a low-dose non-contrast computed tomography attenuation correction (CTAC) scan is often used to correct for soft-tissue attenuation, leading to improved diagnostic accuracy.^3,4^ Attenuation correction by computed tomography (CT) is recommended by American Society of Nuclear Cardiology guidelines.^5^ Although the myocardium is the structure of principal interest during SPECT/CT MPI, its CTAC scan provides a wealth of additional information about other visible organs. Incidental findings have been reported in up to 59.5% of SPECT/CT MPI studies, of which some are clinically important and necessitate further diagnosis and treatment.^6,7^

However, due to limitations in the quality of CTAC images (low dose, no electrocardiographic gating), detection and characterization of abnormal findings on CTAC can be challenging.^8^ Consequently, the additional information present in hybrid cardiac scans is often underutilized during clinical reporting. While some methods have been developed to derive information about coronary artery calcium (CAC) and epicardial adipose tissue (EAT) from CTAC scans, ^9,10^ many other potentially clinically important features, like extracardiac structures, are present in these scans, yet to date their added value to MPI has not been systematically evaluated.

The aim of this study is to develop a holistic artificial intelligence (AI)-based approach for the prediction of all-cause mortality from SPECT/CT MPI utilizing all possible information contained in the hybrid images and to separately evaluate the value of CTAC images for this purpose, which have been previously underutilized.

## 2. Material and methods

### 2.1 Study population

In this retrospective study we utilized CTAC scans of patients who underwent SPECT/CT MPI from 4 sites (University of Calgary, Yale University, Columbia University, University of Ottawa Heart Institute) participating in the Registry of Fast Myocardial Perfusion Imaging with Next generation SPECT (REFINE SPECT).^11^ The study protocol was approved by the IRB at all participating sites and complied with the Declaration of Helsinki. Baseline demographic and clinical characteristics were obtained from the REFINE SPECT registry.^11^ CTAC image acquisition at each participating site is shown in Table S1. The outcome was all-cause mortality (referred to subsequently simply as “mortality”), which was determined using the national death index for sites in the United States and administrative databases in Canada.

### 2.2 Myocardial Perfusion Image Analysis

Total perfusion deficit (TPD), end-diastolic stress shape index (ratio between the maximum left ventricular (LV) diameter in short axis and the length of the LV in end-diastole at stress), stress ejection fraction, and end-diastolic volume were quantified automatically from non-attenuation-corrected MPI scans at the core laboratory (Cedars-Sinai Medical Center, Los Angeles) with the use of dedicated software (Quantitative Perfusion SPECT [QPS] software, Cedars-Sinai Medical Center, Los Angeles)^12^. Normal myocardial perfusion was defined as stress TPD <5%^13^, whereas moderate-to-severe ischemia was defined as TPD ≥10% of the myocardium.^14^

### 2.3 Multi-structure deep learning feature extraction from CTAC

The study design is shown in Figure 1. TotalSegmentator, a multi-structure segmentation deep learning (DL) model, was used to segment structures visible on CTAC.^15^ Out of all segmented structures, we selected thirty-three structures with a frequency of >80% on all scans (Figure S1). The automatic extraction of imaging features for all selected structures was performed with PyRadiomics package (version 3.0.1).^16^ In per-organ analysis, we included eleven first-order and four 3D features (Table S2).

**Figure 1.**
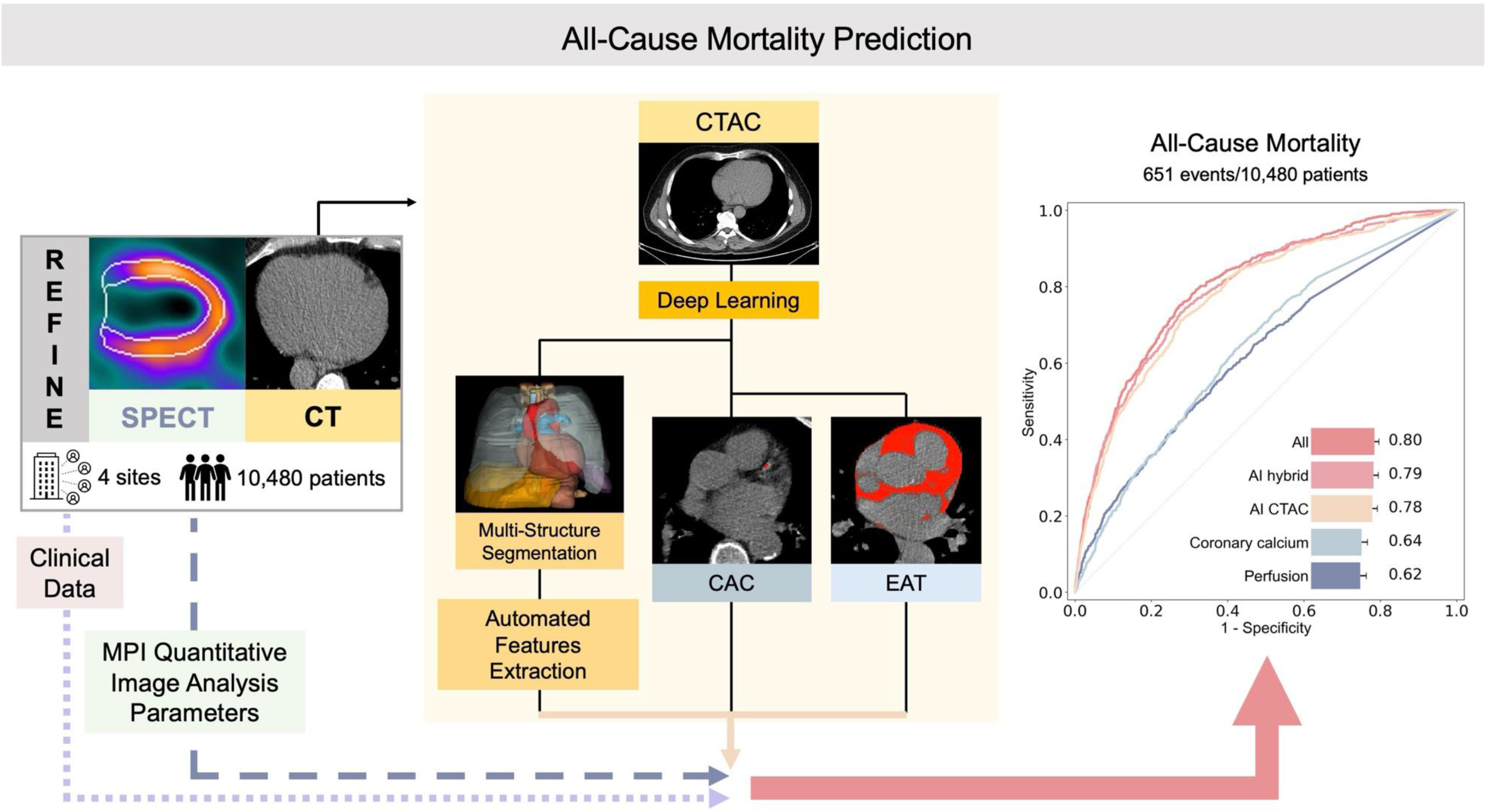
Central illustration. Artificial intelligence (AI) model integrating fully automated multi-structure computed tomography attenuation correction (CTAC) segmentation, quantitative image analysis (radiomics), deep learning (DL)-based coronary artery calcium (CAC), and epicardial adipose tissue (EAT) in all patients undergoing myocardial perfusion imaging (MPI) single-photon emission computed tomography/computed tomography (SPECT/CT). Receiver-operating characteristics curve for all-cause mortality and area under the receiver-operating characteristic curve values of **Coronary calcium** (DL-CAC score), **Perfusion** (stress TPD), the **AI CTAC model** (including DL-CAC, DL-EAT, and radiomics); the **AI hybrid model** – combing the CTAC model with stress MPI quantitative image parameters and stress variables and the **All model** incorporating AI hybrid image features, and clinical data.

### 2.4 Automated Coronary Artery Calcium Scoring

Our formerly validated deep learning model was used for CAC segmentation and scoring.^17,18^ To segment heart mask and CAC on CTAC images, two convolutional long short-term memory (convLSTM) networks were tested externally on data (10,480 CTAC scans) from 4 different sites. To automatically obtain CAC scores from the deep learning segmentation, established methods were used.^19^

### 2.5 Automated Epicardial Adipose Tissue Scoring

A previously developed deep learning model was used to estimate EAT volume and density (-190 and -30 Hounsfield units [HU]) from CTAC scans.^10^ For EAT model training and validation purposes, we used 500 CTAC scans from one site (Yale University). Patients who were used for EAT model training and validation were not included in this analysis.

### 2.6 Classification Models

Extreme Gradient Boosting (XGBoost) models (version 1.7.3), a currently leading machine learning method, were used for mortality classification. These models generate all-cause mortality risk scores by applying 10-fold cross-validation regimen across the entire dataset. Within each fold, 90% of the data was first set aside for model training and validation. This 90% was further divided, with 80% used for training and 20% for validation. The remaining 10% of the data in each fold was used for testing and kept separate from training and validation to ensure each patient was tested exactly once across all folds. 10 separate models were built, and each was tested independently. Testing results were concatenated from all models for the overall performance evaluation. Hyper-parameter tuning to optimize the model parameters was conducted during training and validation, separately in each fold using the grid-search method.

Three key benefits of employing 10-fold cross-validation are primarily 3-fold: 1) it reduces the variability of prediction errors, leading to a more accurate evaluation; 2) it maximizes the data utilization while minimizing the chance of overfitting and cross-contamination of information among data splits; 3) it guards against validating the hypothesis influenced by arbitrary data split (Type III error).^20,21^

### 2.7 Models

Four models were used for the mortality endpoint: 1 – model incorporating DL-EAT (**EAT**), 2 – model combining quantitative CTAC image analysis of all segmented structures [radiomics], DL-EAT and DL-CAC **(AI CTAC**), 3 – model incorporating all variables included in the CTAC model as well as stress ejection fraction, stress end-diastolic volume, stress shape index end-diastolic, stress TPD, and other SPECT imaging features [see Table S3] **(AI hybrid)**, 4 – model combining CTAC, MPI and clinical data (**All)**, whereas **Coronary calcium** (DL-CAC score) and **Perfusion** (utilizing stress TPD) were univariate comparisons.

Clinical data include patient demographics such as age, sex, body mass index [BMI]. Also included is past medical history: hypertension, diabetes, dyslipidemia, prior CAD (prior myocardial infarction, percutaneous coronary intervention [PCI], and coronary artery bypass graft [CABG]). Further, the clinical data encompass variables from stress test such as the type of test, peak stress heart rate, peak stress blood pressure, and ECG response to stress.

### 2.8 Model Explainability

The predictive power of variables included in model training was evaluated using XGBoost feature importance, which quantifies the increase in accuracy resulting from the addition of each feature. SHapley Additive exPlanations (SHAP), a game-theoretic feature importance method, was used to explain how structures contributed to the overall risk in model inference for individual patients.^22^

### 2.9 Thresholds for Comparisons of Machine Learning

Patients were classified into low or high-risk groups based on AI-derived all-cause mortality risk score. This classification was achieved by setting a threshold that aligns with the proportion of patients identified by the established clinical criteria for ischemia (≥10%).^23,24^

### 2.10 Statistical Analysis

Continuous variables with a normal distribution are presented as mean ± standard deviation (SD) and not normally distributed variables as medians with interquartile range (IQR) [IQ1-IQ3]. Categorical variables are expressed as count and relative frequencies (percentages). Differences between categorical variables were compared by the Pearson’s χ2 test whereas continuous variables were compared by Wilcoxon Mann-Whitney test, as appropriate. The performance of the models was evaluated using receiver-operating characteristics analysis, and area under the receiver-operating characteristic (AUC) analysis values were compared with the DeLong test.^25^ Kaplan-Meier survival curve, alongside univariate Cox proportional hazard models, were employed to evaluate the association with mortality. Log-rank test was used to ascertain the statistical significance. The improvement in model predictions was measured using the time-dependent net reclassification improvement score at 2 years.^26^ Confidence intervals were calculated by the percentile bootstrap method. A two-tailed p-value of <0.05 was considered statistically significant. All statistical analyses were performed with Pandas (version 2.1.1) and Numpy (version 1.24.3), Scipy (version 1.11.4), Lifelines (version 0.28.0) and Scikit-learn (version 1.3.0) in Python 3.11.5 (Python Software Foundation, Wilmington, DE, USA), as well as “nricens” package (version 1.6) in R version 4.3.2 (R Foundation for Statistical Computing, Vienna, Austria).

## 3. Results

### 3.1 Patient Characteristics

#### Study population

In total 10,983 participants from 4 sites were enrolled in the REFINE SPECT registry, of which 500 CTAC scans from one site were used for EAT-model training and validation. Of the 10,483 remaining participants, 3 were excluded due to incomplete CTAC scans. The final study cohort consisted of 10,480 participants (Figure S2).

Table 1 represents baseline characteristics stratified by sex. Of all participants, 5,745 (54.8%) were male, and median age was 65 (57, 73) years. During the median 2.9-year (1.6-4.0) follow-up period, 651 (6.2%) patients died. Normal myocardial perfusion was present in 6,165 (58.8%) patients, of whom 274 (4.4%) died. Patients with normal perfusion were significantly younger (p<0.001), more often female, and less often diagnosed with hypertension (p<0.001), diabetes (p<0.001), and dyslipidemia (p=0.007) (Table S4).

**Table 1.**
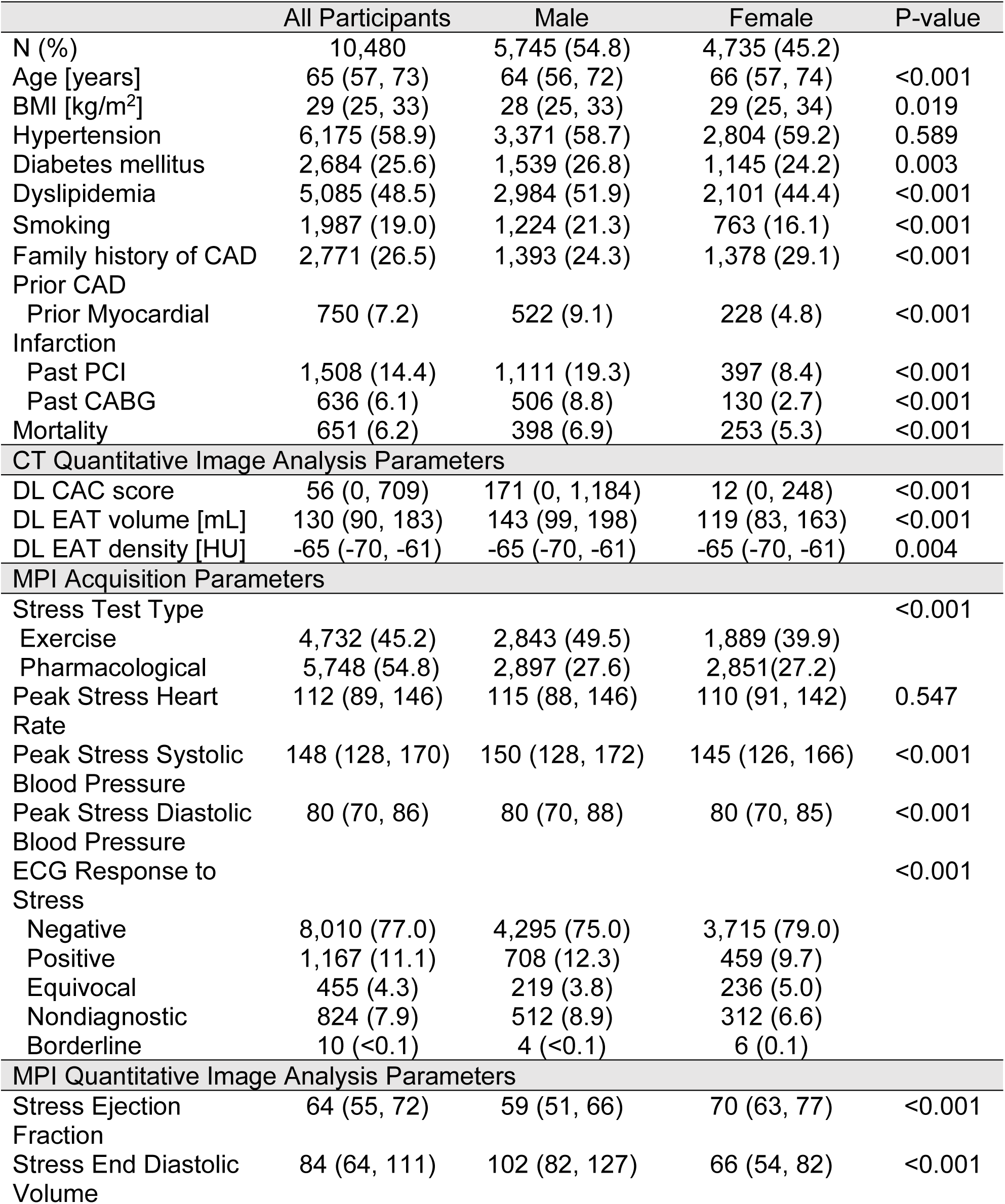
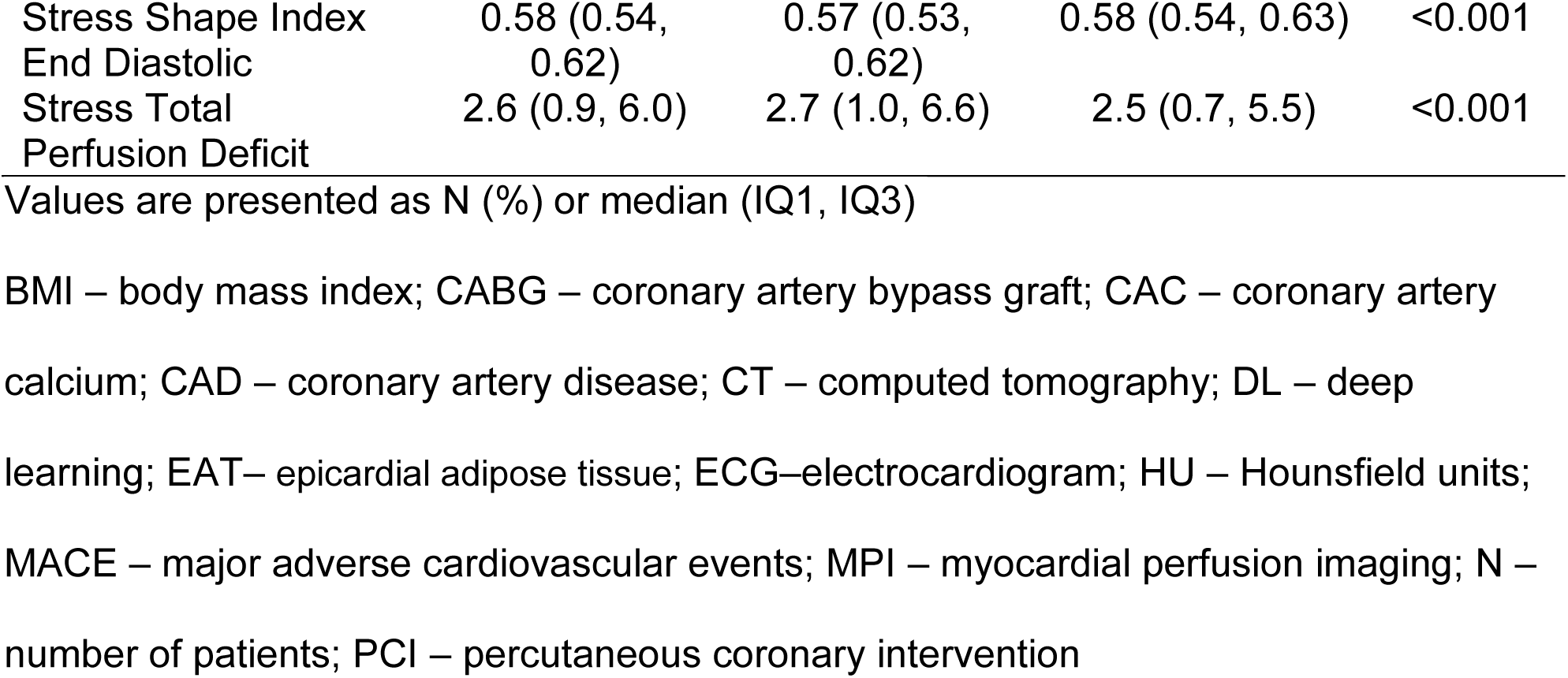
Baseline characteristics for all participants stratified by sex.

#### Myocardial Imaging Perfusion Quantitative Image Analysis Parameters

In all patients, the median total perfusion deficit (TPD) was 2.6% (0.9-6.0) and was higher in male than female patients (2.7 vs. 2.5, respectively, p<0.001) (Table 1). Significantly lower stress ejection fraction was observed in men compared with women (59% vs. 70%, respectively, p<0.001). The median TPD in patients with abnormal perfusion was 7.0 (4.9-11.7), whereas the median stress ejection fraction in this group was 59 (49-68) (Table S4).

#### Coronary Artery Calcium and Epicardial Adipose Tissue

CAC was 0 in 3,753 (35.8%) patients, >0-100 in 1,982 (18.9%), >100-400 in 1,462 (14.0%), and >400 in 3,283 (31.3%) subjects. The median EAT volume and density were 130 mL (90, 183) and -65 HU (-70, -61), respectively (Table 1).

In patients with normal perfusion, 2,459 (39.9%) subjects had no CAC, 1,305 (21.2%) had CAC >0 and ≤100, 862 (14.0%) had CAC >100 and ≤400, and 1,539 (25.0%) had CAC >400. The median EAT volume and density in patients with normal perfusion were 129 mL (89, 179) and -65 HU (-70, -61), respectively (Table S4).

### 3.2 Model Performance

Figure 2 represents the model performance and feature importance for mortality in all patients, subjects with normal perfusion, and patients without calcified lesions in coronary arteries. The lungs were the top feature in all patients, in patients with normal perfusion as well as in subjects without coronary calcifications. Table S5 shows AUCs with 95% CI for all AI models in all patients included in the study. There was a better performance of the AI CTAC model than the EAT model (AUC 0.56, 95% CI 0.49-0.63, p<0.001), and coronary calcium (AUC 0.64, 95% CI 0.57-0.71, p<0.001) alone. There was a statistically significant difference in the prediction performance of the AI hybrid model and the CTAC model (AUC 0.79 vs. 0.78, p<0.001).

**Figure 2.**
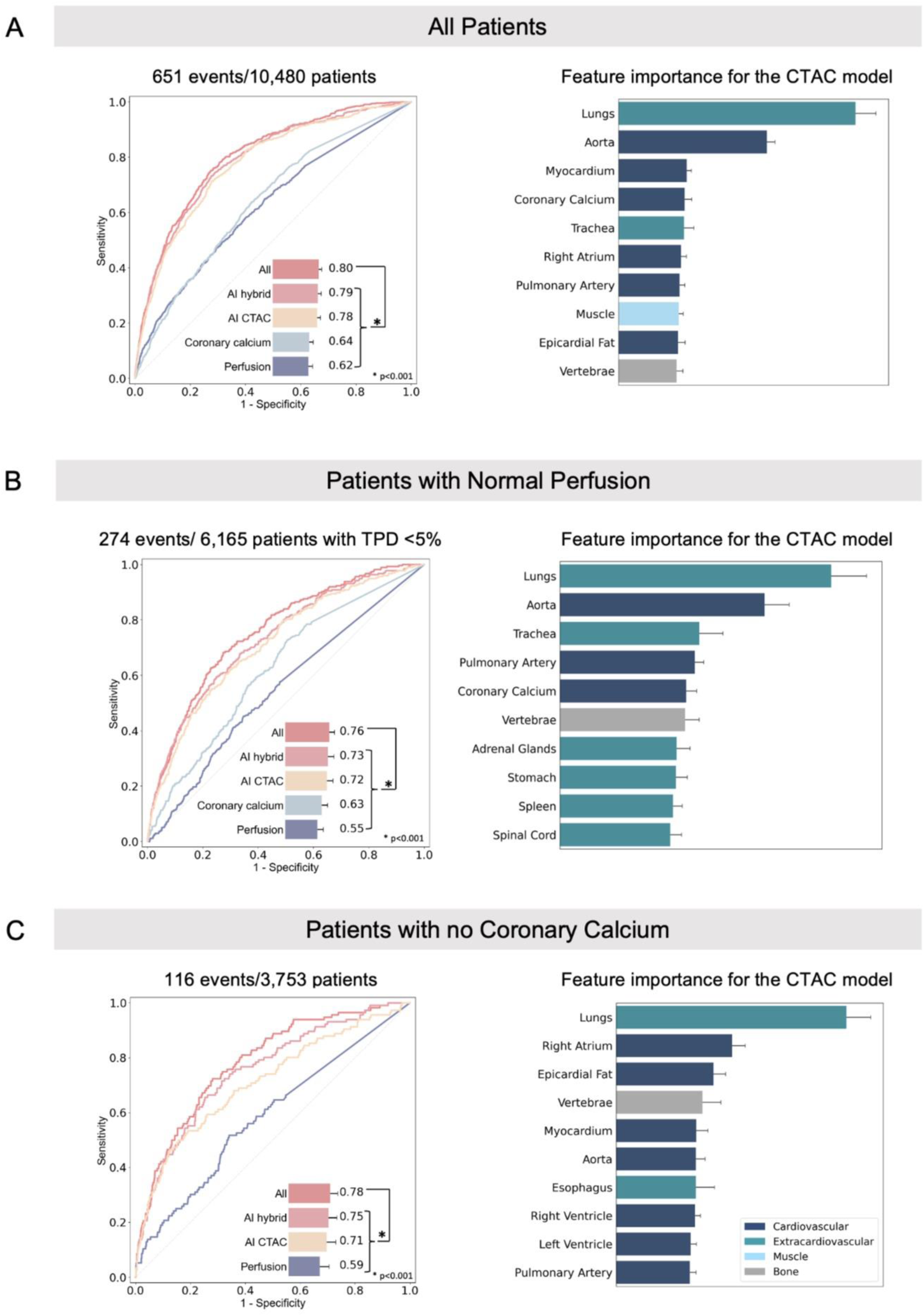
Model performance and feature importance scores for all-cause mortality in all patients (A), in patients with normal perfusion (B), and patients with no coronary artery calcification (C). Normal myocardial perfusion was defined as total perfusion deficit (TPD) <5%. Receiver operating characteristic curve for the **AI computed tomography attenuation correction** (**CTAC**) **model**, including deep-learning (DL) coronary calcium, DL-epicardial adipose tissue (EAT), and radiomics, the **AI hybrid model** incorporating CTAC and myocardial perfusion imaging (MPI) data (stress MPI quantitative image parameters, **Coronary Calcium** (DL-coronary artery calcium score), **Perfusion** (stress TPD), and a model combining CTAC, MPI, and clinical data, (**All**). In all patients, the performance of the EAT model (not shown in the figure) alone was AUC 0.56, in patients with TPD <5% AUC 0.55, whereas in subjects with no coronary calcium AUC 0.59. Feature importance score plot represents 10 segmented structures with the highest scores for the CTAC model.

AUCs with 95% CI for all AI models in patients with normal myocardial perfusion are shown in Table S6 whereas in subjects with no coronary calcium in Table S7. In the group with normal perfusion, the performance of the AI CTAC model was significantly better compared to Perfusion (AUC 0.72 vs. 0.55, respectively, p<0.001). The AI hybrid model incorporating CTAC and MPI features had higher prediction performance compared to the AI CTAC-only model (AUC 0.73 vs. 0.72, respectively, p<0.001). Among the patients with no calcium, the AI CTAC model significantly outperformed Perfusion (AUC 0.71 vs. 0.59, respectively, p<0.001). The AI hybrid model was significantly better than AI CTAC-only model (AUC 0.75 vs 0.71, respectively, p<0.001).

### 3.3 Association with Outcomes and Multivariable Model

Kaplan-Meier Curves stratified by TPD (ischemia <10% and ≥10%), and a matched proportion of patients with high and low AI scores (AI threshold at 0.095, high risk in 9.8%) are shown in Figure 3. AI score led to an improved risk reclassification of patients who experienced mortality (23.9%, 95% CI 19.5-28.5, p<0.001), and patients who did not experience mortality (2.4%, 95% CI 1.7-3.2, p<0.001), with an overall net reclassification improvement of 26.4% (95% CI 21.8-31.0, p<0.001).

**Figure 3.**
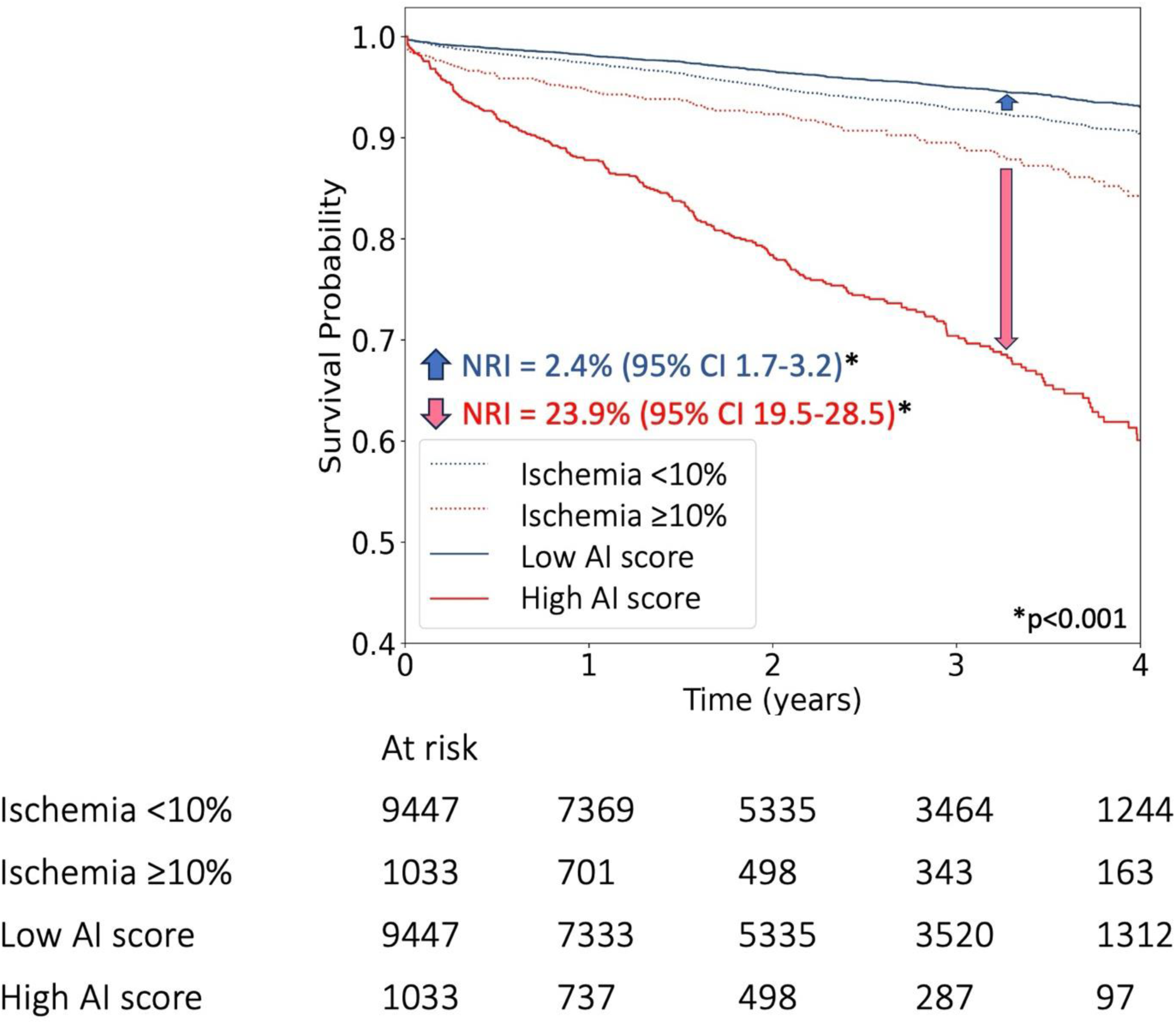
Kaplan-Meier curves stratified by total perfusion deficit (TPD) matched to AI scores (All model). Ischemia was defined as TPD ≥10%. Abbreviations: CI – confidence interval, NRI – Net Reclassification Improvement.

Figure S3 illustrates findings of multivariable analyses. After adjusting for age, sex (male), hypertension, dyslipidemia, diabetes mellitus, peripheral vascular disease, past myocardial infarction, and family history of CAD, patients with abnormal perfusion were at higher risk of death compared to patients with normal myocardial perfusion (adjusted hazard ratio [HR] 1.71, 95% CI 1.46-2.01, p<0.001). Moreover, CAC >400 (adjusted HR 2.11, 95% CI 1.67-2.65, p<0.001) was associated with an increased risk of death.

### 3.4 Structure Specific Risk Evaluation

Examples of patients classified to be at a higher risk of death (with extracardiac structures, notably the lungs and aorta, contributing the most to mortality) are shown in Figure 4.

**Figure 4.**
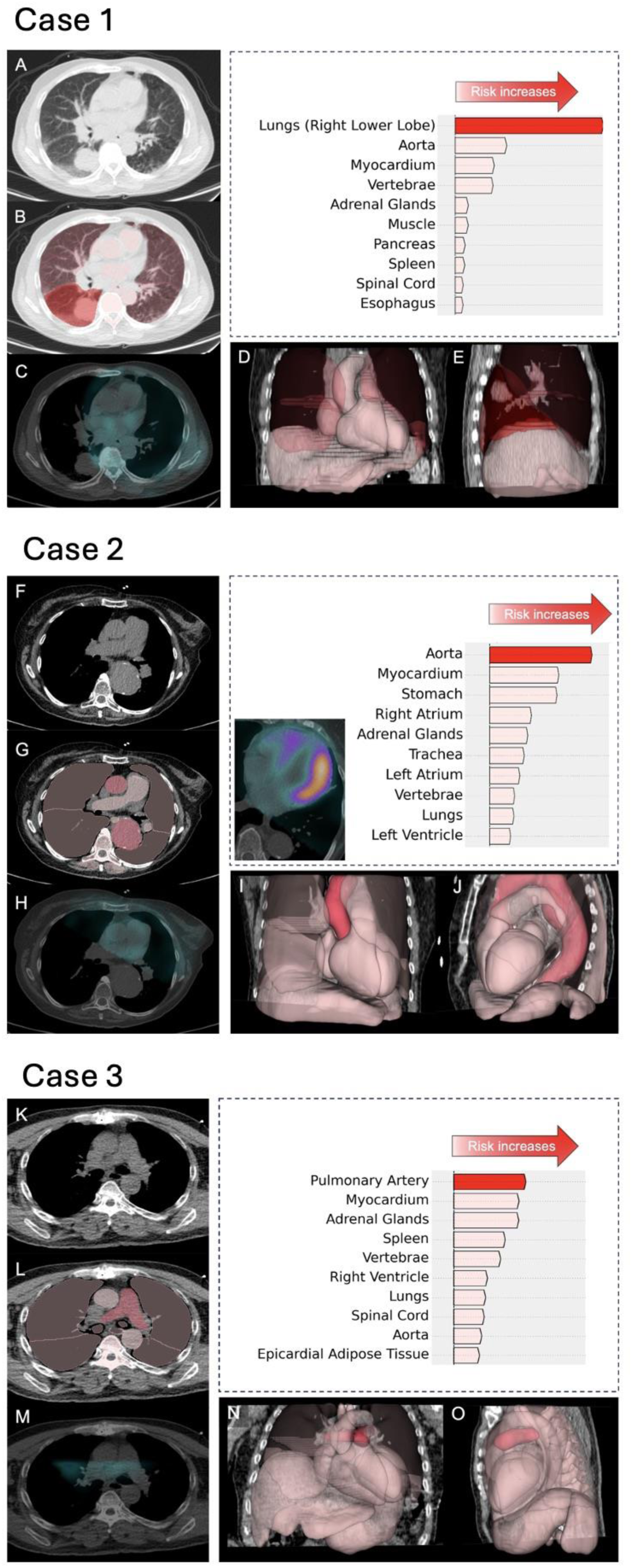
Examples of patients undergoing single-photon emission computed tomography/computed tomography (SPECT/CT) myocardial perfusion imaging with an extracardiac structure increasing the highest risk of all-cause mortality. Waterfall plot demonstrates the top 10 structures with the highest influence on per-patient risk of mortality (for the computed tomography attenuation correction [CTCA] model). **Case 1**: A male patient was classified to be at higher risk of death with the lungs (right lower lobe) contributing the most to the elevated risk (red arrow on the waterfall plot). **A.** CTAC, axial view, with a corresponding deep learning (DL) structures segmentation (**B**) revealed a 39x39 mm solid mass with irregular margins in the right lower lobe. CTAC with an overlayed SPECT scan showed no uptake of the radiotracer. **D-E.** 3D reconstruction of all segmented and ranked structures. The patient had abnormal myocardial perfusion (total perfusion deficit of 7.65) and died 48 days after the exam. **Case 2:** A male patient was identified to be at elevated risk of mortality. The risk of death was increased the most by the aorta (red arrow on the waterfall plot). **F**. CTAC, axial view, with a corresponding DL structures segmentation (**G**) showed a descending aortic aneurysm (cross-sectional diameters of the descending aorta 53x51 mm at the level of the pulmonary artery bifurcation). **H**. CTAC with an overlayed single-photon emission tomography scan, coronal view. **I-J**. 3D reconstruction of all segmented and ranked structures. The patient had abnormal myocardial perfusion (total perfusion deficit of 32.1) and died 209 days after the exam. **Case 3:** A male patient was assigned to be at increased risk of death. The dilated pulmonary artery was contributing the most to the elevated risk (red arrow on the waterfall plot). **K**. CTAC with a corresponding DL structures segmentation (**L**) showed a dilatated pulmonary artery (the pulmonary artery diameter on transaxial image - 32 mm). **M**. CTAC with an overlayed SPECT scan showed no uptake of the radiotracer. **N-O**. 3D reconstruction of all segmented and ranked structures. The patient had abnormal myocardial perfusion (total perfusion deficit of 9.67) and died 5.4 years after the exam.

## 4. Discussion

In this study, we have demonstrated the potential value of holistic anatomic, functional and clinical evaluation of CTAC scans for improving all-cause mortality prediction in patients undergoing hybrid perfusion MPI. We developed a fully automated AI model incorporating multi-structure segmentation and radiomic feature extraction in parallel to deep learning-based CAC and EAT quantification. This model improves mortality prediction from multimodality myocardial perfusion, with a combined model improving upon any feature set (SPECT, CTAC, or clinical) in isolation. Moreover, it provides physicians with guidance regarding portions of CTAC scans which require further scrutiny to identify potentially important underlying conditions indicating potentially significant incidental findings, despite coronary artery disease being the primary indication for the examination. This fully automated workflow could be leveraged by physicians to unlock the full potential of hybrid SPECT/CT imaging.

Several studies have proven the role of AI in predicting cardiovascular events from SPECT/CT using clinical, MPI ^27,28^, and CTAC data.^29,30^ Nevertheless, only a limited number of CTAC findings, like CAC^29^, or EAT^10^ were included in these previous analyses. More recently we demonstrated that deep learning cardiac chamber volumes (from CTAC) provided incremental and complementary value to CAC and SPECT variables.^31^ Ashrafinia et al. used radiomic features from SPECT MPI to predict CAC score derived from CT scans^32^, whereas Amini et al. applied a quantitative image analysis approach not only to diagnose CAD, but also for risk classification.^33^ The proposed AI approach integrates simultaneous assessment of multiple structures on CTAC by leveraging strengths of deep learning and quantitative image analyses. Importantly, the model incorporating SPECT, CTAC, and clinical data had the highest prediction performance suggesting that AI-derived information encrypted in CTAC is complementary to traditional methods for analysis.

By integrating functional imaging (SPECT) with anatomic characteristics (CT), hybrid imaging has not only enhanced nuclear medicine by improving diagnostic accuracy ^34,35^, but also provides an enormous amount of data contained in CTACs - which to date is not fully utilized. However, accurate interpretation and identification can be challenging due to image quality of these low-dose, non-electrocardiographically gated, and often free- breathing scans.^8^ Importantly, these auxiliary scans may be interpreted by physicians without dedicated training in interpreting chest CT.^36^ In some cases, unexpected intra-, and extrathoracic radiotracer uptake can lead to identification of conditions like thymoma, breast and lung cancer.^37,38^ Previous studies have demonstrated the ability of AI to support physicians in identifying potentially important incidental findings.^39,40^ Our AI model could potentially help with this clinical challenge by combining 33 cardiac and extracardiac structures automatically segmented from CTAC scans, and ranking those structures based on their importance in predicting mortality in each patient.

### Limitations

This study has some limitations. It was a retrospective study with non-uniform CTAC acquisition protocols from multiple sites, however, this highlights the generalizability of the approach. Some organs were only partially visible or not visualized on all scans. For example, organs like kidneys and thyroid were excluded from the analysis because of their high missingness across the cohort. No information regarding the reported cause of death is available in this large, multicenter registry. Therefore, we are not able to evaluate the associations between organ features and cause-specific death. Finally, radiological evaluation of CTACs was performed only with radiomic features and no information regarding reported incidental findings is available in this cohort.

### Conclusions

We demonstrate a significant, yet underappreciated, role of CTAC in risk stratification with MPI SPECT/CT. Fully automated AI integration of quantitative features from multiple organs derived from CTAC, perfusion and clinical data images significantly improves mortality risk stratification in patients undergoing SPECT/CT MPI as compared to MPI only.

## Data Availability

To the extent allowed by data sharing agreements and IRB protocols, the data from this manuscript will be shared upon written request.

## Acknowledgements

This research was supported in part by grants R01HL089765 and R35HL161195 from the National Heart, Lung, and Blood Institute at the National Institutes of Health (PI: Piotr Slomka). The content is solely the responsibility of the authors and does not necessarily represent the official views of the National Institutes of Health. MB is supported by a research award from the Kosciuszko Foundation – The American Centre of Polish Culture.

## List of abbreviations

AI: artificial intelligence
AUC: area under the receiver-operating characteristic curve
BMI: body mass index
CABG: coronary artery bypass graft
CAC: coronary artery calcium
CAD: coronary artery disease
CI: confidence interval
convLSTM: convolutional long short-term memory
CT: computed tomography
CTAC: computed tomography attenuation correction
DL: deep learning
EAT: epicardial adipose tissue
HR: hazard ratio
HU: Hounsfield units
LVEF: left ventricular ejection fraction
MPI: myocardial perfusion imaging
PCI: percutaneous coronary intervention
SPECT: single-photon emission computed tomography
TPD: total perfusion deficit

