## Supplemental Material for "Holistic AI analysis of hybrid cardiac perfusion images for mortality prediction"

**sSUPPLEMENTARY MATERIAL**

Table S1	Page 2

Table S2	Page 3

Table S3	Page 4

Table S4	Page 5-6

Table S5	Page 7

Table S6	Page 8

Table S7	Page 9

Figure S1	Page 10

Figure S2	Page 11

Figure S3	Page 12-13

**Table S1**. Computed tomography attenuation correction image acquisition parameters

| Site | University of Calgary | Yale University | Columbia | Ottawa |
| --- | --- | --- | --- | --- |
| Scanner | GE Discovery NM/CT 570c | GE Discovery NM/CT 570c | Phillips Precedence 16P | Siemens Symbia Intevo 16 |
| ECG-gating | No | No | No | No |
| Breath-holding | Yes (end expiratory) | No | No | Yes (end-expiratory) |
| Slice Thickness [mm] | 5.0 | 2.5 | 3.0 | 5.0 |
| Tube Current [mA] | 16 | 60/150* | 30 | 20 |
| Tube Voltage [kVp] | 120 | 120 | 120 | 120 |

*acquisition parameters adjusted for patients with body mass index >= 40 kg/m^2^

**Table S2.** Selected organs, radiomics features and MPI parameters

| Organs (n=33) | First-Order Features^‡^ (n=11) | 3D Shape Features^‡^ (n=4) |
| --- | --- | --- |
| Myocardium | 10^th^ Percentile | Elongation |
| Left Atrium | 90^th^ Percentile | Flatness |
| Left Ventricle | Kurtosis | Sphericity |
| Right Atrium | Maximum | Voxel Volume |
| Right Ventricle | Minimum |  |
| Left Atrial Appendage | Mean |  |
| Aorta | Robust Mean Absolute Deviation |  |
| Pulmonary Artery | Root Mean Squared |  |
| Lungs^**^  Trachea | Skewness  Total Energy |  |
| Esophagus | Uniformity |  |
| Stomach |  |  |
| Liver |  |  |
| Spleen |  |  |
| Pancreas |  |  |
| Adrenal Glands^*^ |  |  |
| 4^th^ - 12^th^ Thoracic Vertebrae |  |  |
| Erector Spinae Muscle^*^ |  |  |
| Spinal cord |  |  |

^*^ right and left, ^**^ right upper lobe, right middle lobe, right lower lobe, left upper lobe, left lower lobe

^‡^ Radiomics

MPI – myocardial perfusion imaging

**Table S3.** SPECT imaging features

|  | Features |
| --- | --- |
| Stress | Total Perfusion Deficit |
|  | Quality control |
|  | Volume |
|  | Shape index, end-diastolic |
|  | Shape index, end-systolic |
|  | Length |
| Stress gated | Ejection fraction |
|  | Volume, end-diastolic |
|  | Quality control |
|  | Shape index, end-diastolic |
|  | Shape index, end-systolic |
|  | Length |
|  | Motion extent for all segments |
|  | Motion raw for all segments |
|  | Thickening extent for all segments |
|  | Thickening raw for all segments |
|  | Wall volume, end-diastolic |
|  | Wall volume, end-systolic |
|  | Volume, end-systolic |
|  | Band width for the entire ventricular counts in the segment |
| Rest | Total perfusion Deficit |

SPECT – single-photon emission computed tomography

**Table S4.** Baseline characteristics for all participants stratified by TPD

|  | All Participants | Abnormal Perfusion | Normal Perfusion | P-value |
| --- | --- | --- | --- | --- |
| N (%) | 10,480 | 4,315 (41.2) | 6,165 (58.8) |  |
| Age [years] | 65 (57, 73) | 66 (57, 74) | 64 (56, 72) | <0.001 |
| Male | 5,745 (55) | 2,442 (57) | 3,303 (53.6) | 0.002 |
| BMI [kg/m^2^] | 29 (25, 33) | 29 (25, 34) | 28 (25, 33) | <0.001 |
| Hypertension | 6,175 (58.9) | 2,673 (61.9) | 3,502 (56.8) | <0.001 |
| Diabetes mellitus | 2,681 (25.6) | 1,322 (30.7) | 1,359 (22.0) | <0.001 |
| Dyslipidemia | 5,085 (48.5) | 2,162 (50.1) | 2,923 (47.4) | 0.007 |
| Smoking | 1,987 (19.0) | 773 (17.9) | 1,214 (19.7) | 0.024 |
| Family history of CAD | 2,771 (26.4) | 1,078 (24.9) | 1,693 (27.5) | 0.005 |
| Prior CAD |  |  |  |  |
| Prior Myocardial Infarction | 750 (7.2) | 465 (10.8) | 285 (4.6) | <0.001 |
| Past PCI | 1,508 (14) | 888 (20.6) | 620 (10.1) | <0.001 |
| Past CABG | 636 (6.1) | 444 (10.3) | 192 (3.1) | <0.001 |
| Mortality | 651 (6.2) | 377 (8.7) | 274 (4.4) | <0.001 |
| CT Quantitative Image Analysis Parameters | | | | |
| DL-CAC score | 56 (0, 709) | 166 (0, 1,175) | 25 (0, 399) | <0.001 |
| DL-EAT volume [mL] | 130 (90, 183) | 133 (92, 189) | 129 (89, 179) | 0.003 |
| DL-EAT density [HU] | -65 (-70, -61) | -65 (-70, -61) | -65 (-70, -61) | 0.017 |
| MPI Acquisition Parameters |  |  |  |  |
| Stress Test Type |  |  |  | <0.001 |
| Exercise | 4,732 (45.2) | 1,685 (39.0) | 3,047 (49.4) |  |
| Pharmacological | 5,748 (54.8) | 2,630 (61.0) | 3,118 (50.6) |  |
| Peak Stress Heart Rate | 112 (89, 146) | 104 (85, 137) | 120 (92, 150) | <0.001 |
| Peak Stress Systolic Blood Pressure | 148 (128, 170) | 141 (122, 164) | 150 (130, 172) | <0.001 |
| Peak Stress Diastolic Blood Pressure | 80 (70, 86) | 78 (70, 84) | 80 (70, 88) | <0.001 |
| ECG Response to Stress |  |  |  | <0.001 |
| Negative | 8,010 (76.4) | 3,175 (73.6) | 4,835 (78.4) |  |
| Positive | 1,167 (11.1) | 537 (12.4) | 630 (10.2) |  |
| Equivocal | 455 (4.3) | 162 (3.8) | 293 (4.8) |  |
| Nondiagnostic | 824 (7.9) | 433 (10.0) | 391 (6.4) |  |
| Borderline | 10 (<0.1) | 3 (<0.1) | 7 (<0.1) |  |
| MPI Quantitative Image Analysis Parameters | | | |  |
| Stress Ejection Fraction | 64 (55, 72) | 59 (49, 68) | 66 (59, 73) | <0.001 |
| Stress End Diastolic Volume | 84 (64, 111) | 93 (69, 126) | 79 (62, 103) | <0.001 |
| Stress Shape Index End Diastolic | 0.58 (0.54, 0.62) | 0.59 (0.54, 0.65) | 0.57 (0.53, 0.61) | <0.001 |
| Stress Total Perfusion Deficit | 2.6 (0.9, 6.0) | 7.0 (4.9, 11.7) | 1.1 (0.3, 2.1) | <0.001 |

Values are presented as N (%) or median (IQ1, IQ3); Normal perfusion – TPD <5%

BMI – body mass index; CABG – coronary artery bypass graft; CAC – coronary artery calcium; CAD – coronary artery disease; CT – computed tomography; DL – deep learning; EAT– epicardial adipose tissue; ECG–electrocardiogram; HU – Hounsfield units; MACE – major adverse cardiovascular events; MPI – myocardial perfusion imaging; N – number of patients; PCI – percutaneous coronary intervention; TPD – total perfusion deficit

**Table S5.** Area under the receiver-operating characteristic curve (AUC) with 95% confidence interval (CI) for all artificial intelligence models and coronary calcium and perfusion (stress total perfusion deficit) in all patients.

| Model | AUC | 95% CI | P-value |
| --- | --- | --- | --- |
| All | 0.80 | 0.74-0.87 | Reference |
| AI hybrid | 0.79 | 0.72-0.86 | <0.001 |
| AI CTAC | 0.78 | 0.71-0.85 | <0.001 |
| Coronary Calcium | 0.64 | 0.57-0.71 | <0.001 |
| Perfusion | 0.62 | 0.55-0.70 | <0.001 |
| EAT | 0.56 | 0.49-0.63 | <0.001 |

The models are described in the Methods section.

AI – artificial intelligence; CTAC – computed tomography attenuation correction; EAT – epicardial adipose tissue.

**Table S6.** Area under the receiver-operating characteristic curve (AUC) with 95% confidence interval (CI) for all artificial intelligence models, coronary calcium and perfusion (stress total perfusion deficit) in patients with normal myocardial perfusion.

| Model | AUC | 95% CI | P-value |
| --- | --- | --- | --- |
| All | 0.76 | 0.65-0.86 | Reference |
| AI hybrid | 0.73 | 0.62-0.84 | <0.001 |
| AI CTAC | 0.72 | 0.61-0.83 | <0.001 |
| Coronary Calcium | 0.63 | 0.51-0.74 | <0.001 |
| EAT | 0.55 | 0.44-0.66 | <0.001 |
| Perfusion | 0.55 | 0.44-0.66 | <0.001 |

The models are described in the Methods section.

AI – artificial intelligence; CTAC – computed tomography attenuation correction; EAT – epicardial adipose tissue.

**Table S7.** Area under the receiver-operating characteristic curve (AUC) with 95% confidence interval (CI) for all artificial intelligence models, coronary calcium and perfusion (stress total perfusion deficit) in patients with no coronary calcium.

| Model | AUC | 95% CI | P-value |
| --- | --- | --- | --- |
| All | 0.78 | 0.63-0.94 | Reference |
| AI hybrid | 0.75 | 0.60-0.92 | <0.001 |
| AI CTAC | 0.71 | 0.55-0.88 | <0.001 |
| EAT | 0.59 | 0.42-0.76 | <0.001 |
| Perfusion | 0.59 | 0.41-0.76 | <0.001 |
| Coronary Calcium | 0.50 | 0.33-0.67 | <0.001 |

The models are described in the Methods section.

AI – artificial intelligence; CTAC – computed tomography attenuation correction; EAT – epicardial adipose tissue.

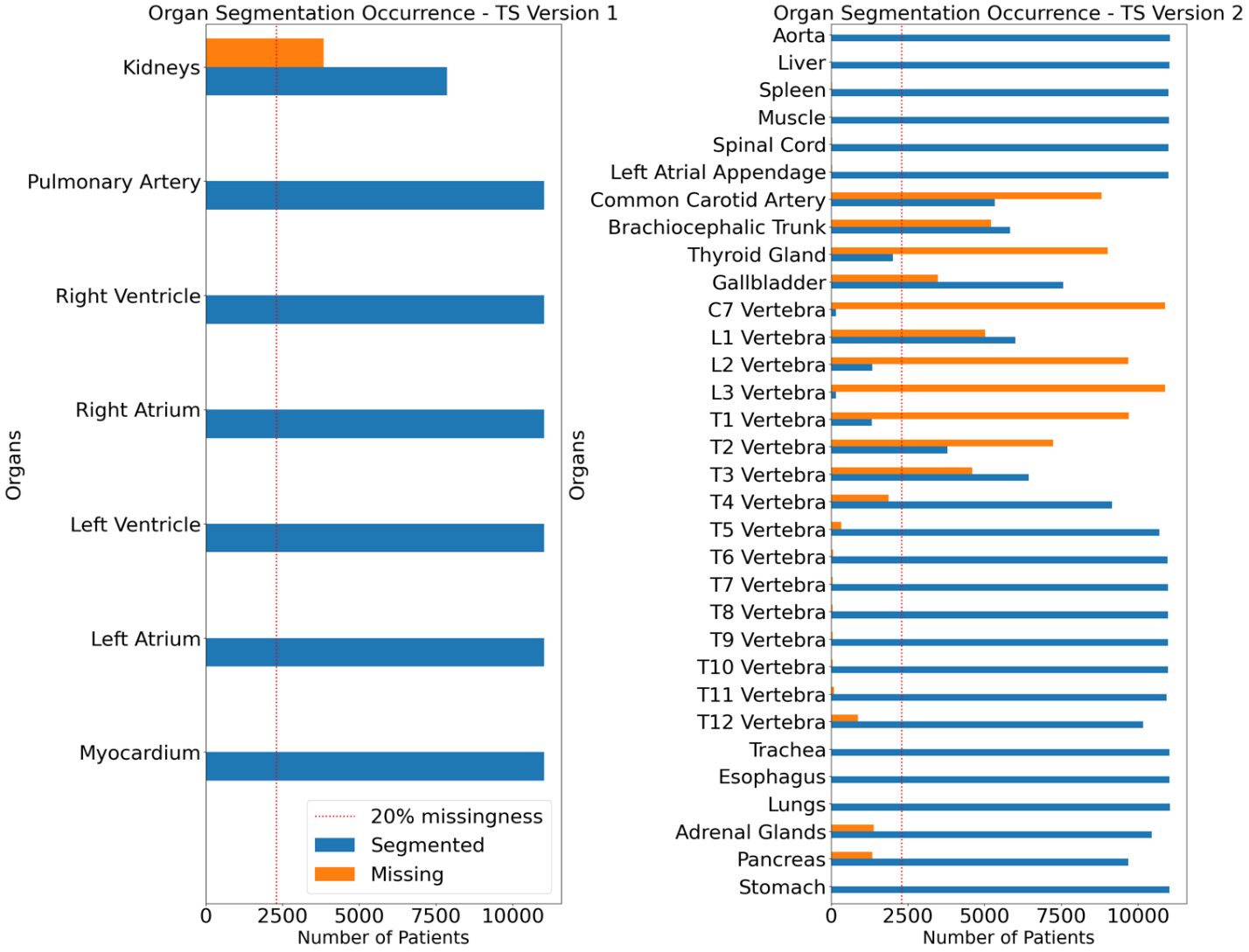

**Figure S1.** Histogram of successfully segmented structures visible on computed tomography attenuation correction (CTAC) scans with a use of deep-learning models. Structures present in >80% of CTAC were included. Abbreviations: TS – TotalSegmentator.

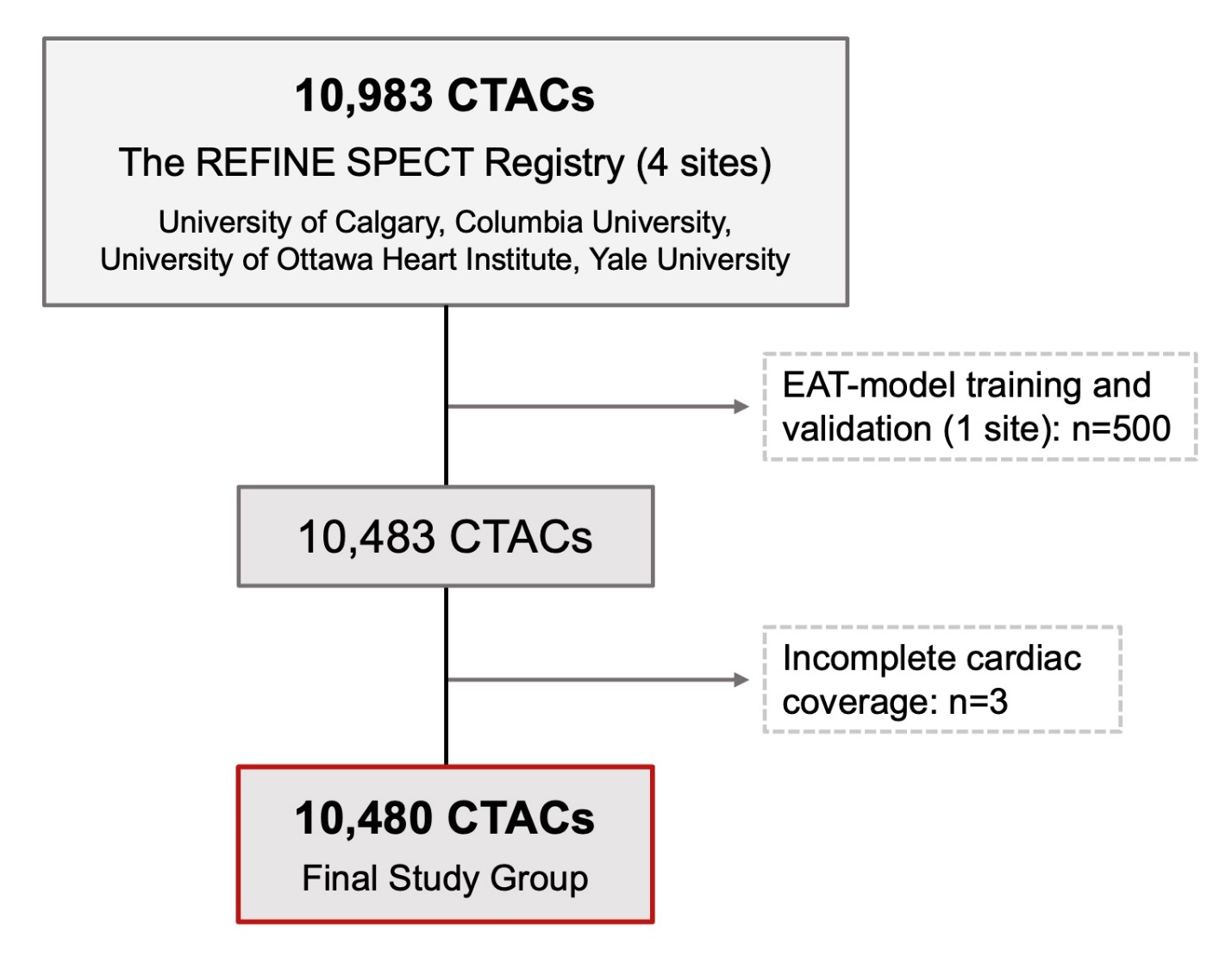

**Figure S2.** Flowchart. Abbreviations: CTAC – computed tomography attenuation correction; EAT – epicardial adipose tissue; SPECT – single-photon emission computed tomography.

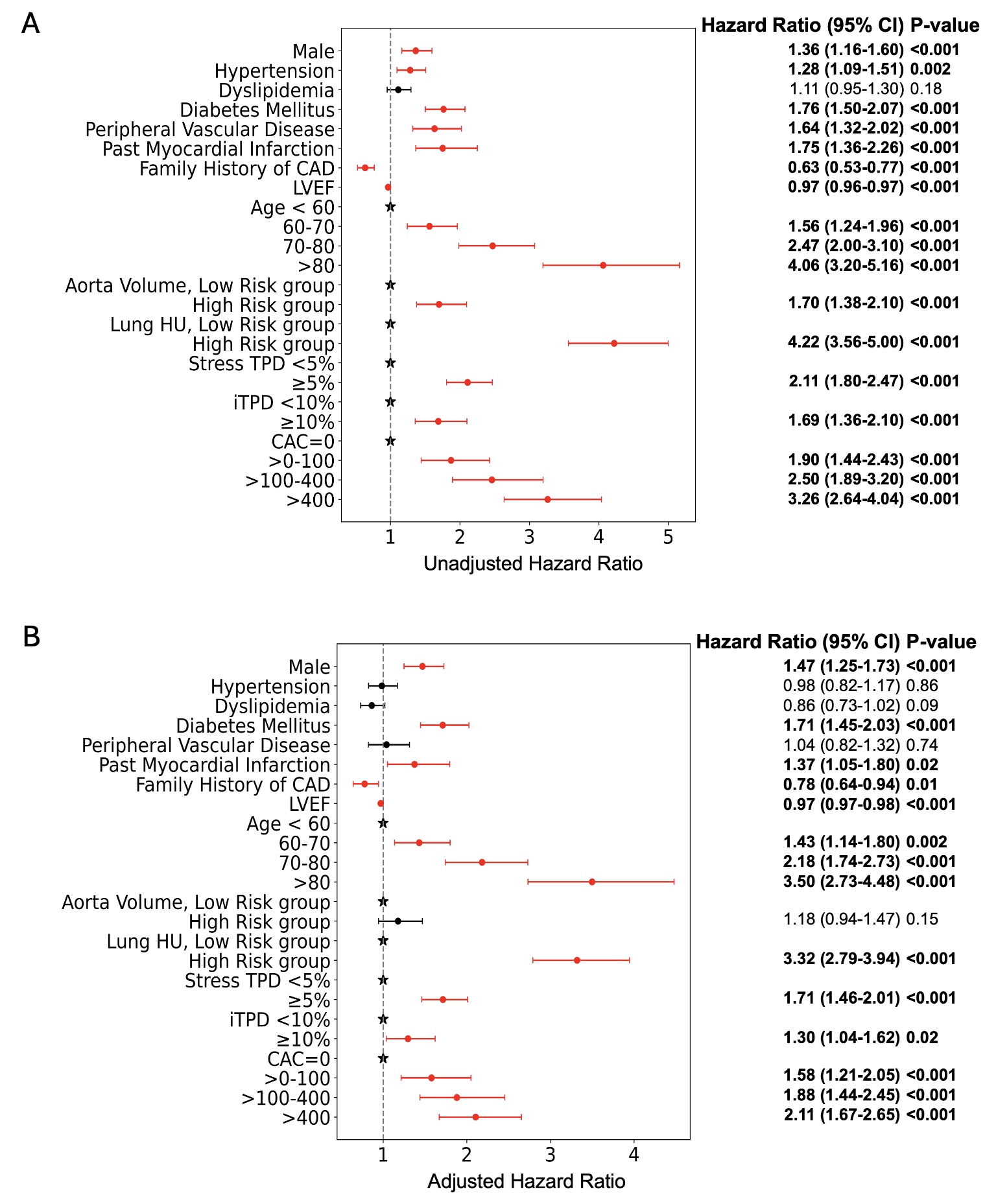

**Figure S3.** Forest plot of the unadjusted (A) and adjusted (B) hazard ratio and 95% confidence interval (CI) for death. Adjusted factors: age, sex (male), hypertension, dyslipidemia, diabetes mellitus, peripheral vascular disease, past myocardial infarction, family history of coronary artery disease (CAD). Abbreviations: CAC – coronary artery calcium, HU – Hounsfield units, iTPD – ischemic total perfusion deficit, LVEF – left ventricular ejection fraction, TPD – total perfusion deficit.
